# Does healthy lifestyle attenuate the detrimental effects of urinary polycyclic aromatic hydrocarbons on phenotypic aging? An analysis from NHANES 2001–2010

**DOI:** 10.1101/2021.12.17.21268014

**Authors:** Zhenqing Yang, Fan Pu, Xingqi Cao, Xueqin Li, Sudan Sun, Jingyun Zhang, Chen Chen, Liyuan Han, Yongli Yang, Wei Wang, Yawei Zhang, Zuyun Liu

## Abstract

**Objectives:** It remains unknown whether healthy lifestyle attenuates the detrimental effects of urinary monohydroxy polycyclic aromatic hydrocarbons (OH-PAHs) on aging. This study aims: 1) to examine the associations of OH-PAHs and lifestyle with Phenotypic Age Acceleration (PhenoAge.Accel), a novel aging measure, respectively; and 2) to evaluate the potential interaction effects of OH-PAHs and lifestyle on PhenoAge.Accel.

**Methods:** Cross-sectional data of 2579 participants (aged 20-84 years, n=1292 females) from the National Health and Nutrition Examination Survey for years 2001-2010 were analyzed. A lifestyle index was constructed based on five components (drinking, smoking, body mass index, physical activity, and diet), ranging from 0 to 5. We calculated PhenoAge.Accel using algorithms developed previously. General linear regression models were used to examine the associations.

**Results:** We observed strong associations of OH-PAHs and lifestyle with PhenoAge.Accel. For instance, one unit increase in ∑NAP (sum of 1- and 2-hydroxynaphthalene) was associated with 0.37 year (95% confidence interval [CI]: 0.26, 0.48) increase in PhenoAge.Accel. We did not observe statistically significant interaction effects between OH-PAHs and lifestyle on PhenoAge.Accel. After stratified by sex, we observed strong associations as well as statistically significant interactions of OH-PAHs and lifestyle with PhenoAge.Accel among females.

**Conclusions:** Both OH-PAHs and lifestyle were independently associated with phenotypic aging and there were statistically significant interactions between OH-PAHs and lifestyle on phenotypic aging among females. The findings highlight the importance of adherence to a healthy lifestyle to attenuate the detrimental effects of exposures to PAHs on phenotypic aging among females.

## 1. Introduction

Polycyclic aromatic hydrocarbons (PAHs) are pivotal components of air pollutants, as they are produced by incomplete combustion of organic materials such as tobacco, fossil fuels, and even through cooking (Kim et al. 2013). Multiple potential pathways (e.g., dermal absorption (McClean et al. 2004), ingestion (Veyrand et al. 2013), inhalation (Zhang et al. 2014)) have led to widespread general population exposure to PAHs. PAHs are metabolized through the liver and excreted in urine and feces with an average half-life of < 30h in the human body (Buckley and Lioy 1992; Motorykin et al. 2015). Because of the short half-life of PAHs, many metabolites in urinary samples have been identified as proper biomarkers of recent exposure to PAHs (Grainger et al. 2006). Existing evidence has shown that exposure to PAHs increases risk of many chronic diseases, including cardiovascular disease (Burstyn et al. 2005), diabetes (Stallings-Smith et al. 2018; Wang et al. 2019), and respiratory illness (Abdel-Shafy and Mansour 2016).

Aging is a critical risk factor for many chronic diseases (Kennedy et al. 2014). As a complex process, aging represents changes in body composition, homeostatic mechanisms, energetics, and brain health over time (Ferrucci et al. 2018), which could be reflected by multisystem clinical chemistry biomarkers. We have recently developed a novel aging measure, Phenotypic Age Acceleration (PhenoAge.Accel), derived from nine chemistry biomarkers (Levine et al. 2018; Liu et al. 2018). PhenoAge.Accel has been demonstrated to capture morbidity and mortality risk across diverse subpopulations in the US (Liu et al. 2018). Given the close connection between aging and chronic diseases, it is intuitive to speculate that PAHs increase the risk of chronic diseases partially through accelerating the phenotypic aging process, which is in line with the Geroscience paradigm (Kennedy et al. 2014).

Of note, aging is strongly responsive to many other risk factors, e.g., modifiable lifestyle factors. We have shown that a healthy lifestyle index consisting of non-drinking, a healthy diet, doing exercise, non-smoking and healthy body mass index (BMI) was significantly associated with an aging measure derived from chemistry biomarkers in Chinese oldest-old population (Jin et al. 2021). Together with other studies (Astuti et al. 2017; Robinson et al. 2020), we highlight the importance of adherence to a healthy lifestyle to slow aging in daily life. Hence, it is intriguing to examine the role of lifestyle in the potential associations between PAHs and phenotypic aging. We hypothesize that there would be an interaction effect of PAHs and lifestyle on phenotypic aging. In other words, the strength of the detrimental effects of PAHs on phenotypic aging would be attenuated in persons with a healthy lifestyle.

Therefore, we analyzed data from the National Health and Nutritional Examination Surveys in 2001-2010 (NHANES 2001-2010) to examine the associations of PAHs and lifestyle with PhenoAge.Accel, a novel aging measure; and to evaluate the potential interaction effects of PAHs and lifestyle on PhenoAge.Accel.

## 2. Materials and Methods

### 2.1. Study Population

Briefly, NHANES is a program designed to assess the health and nutritional status of civilian in the US, using a complex, multistage sampling procedure. Through examinations in a Mobile Exam Center (MEC) and at-home interviews, NHANES collected a large amount of information (e.g., demographics, socioeconomic status, dietary, and health-related questions, and medical and physiological measurements) from a nationally representative sample biennially in counties across the country. Details of the procedures, study design and recruitment for NHANES are provided by the Centers for Disease Control and Prevention (CDC) (NHANES website: https://www.cdc.gov/nchs/nhanes/index.htm). The protocol of NHANES was approved by the National Center for Health Statistics Research Board.

Five waves of NHANES (2001-2002, 2003-2004, 2005-2006, 2007-2008, 2009-2010, with information on both PAHs and PhenoAge) were combined and used in this study (N = 52195). Participants with missing data on blood biomarkers for calculating PhenoAge (who did not provide blood samples, N=42488), on education level (N = 11), poverty income ratio (PIR, N = 666), and urinary creatinine (Cr, N = 70) or did not provide urine samples for testing urinary PAHs (N = 6250) were excluded. For each PAH, we excluded the outliers whose values (after correction by creatinine) were higher than mean + 3 standard deviation (SD) or lower than mean – 3 SD. The final analytic sample included 2579 participants aged 20-84 years (Figure S1).

### 2.2. Measurements of OH-PAHs and creatinine in urine

Our analyses included eight urinary monohydroxy polycyclic aromatic hydrocarbons (OH-PAHs) and created five summarized urinary PAH measurements through grouping them by parent PAH (Grainger et al. 2006): Naphthalene (1- and 2-hydroxynaphthalene, ∑NAP); Fluorene (2- and 3-hydroxyfluorene, ∑FLU); Phenanthrene (1-, 2-, and 3-hydroxyphenanthrene, ∑PHE); Pyrene (1-hydroxypyrene, ∑PYR) and ∑OH-PAH (sum of eight OH-PAHs). Details of sample collection and measurements were described elsewhere (NHANES website). In brief, participants’ spot urinary specimen was collected at MEC, stored at -20°C and then transferred to CDC laboratory for analysis. After enzymatic hydrolysis and solid-phase extraction, OH-PAHs were analyzed through isolate dilution capillary gas chromatography combined with mass spectrometry. The percentage of OH-PAHs above the analytical limit of detection (LOD) was > 98% in NHANES 2001-2010 waves. Considering the majority of the LODs for OH-PAHs vary across NHANES waves, we utilized the method provided by the CDC and assigned the maximal LOD divided by the square root of two to any PAH below the corresponding LOD in each wave. The LOD of each PAH is published in the Laboratory Procedure Manual for each NHANES wave (Brockmeyer et al. 2018; Fan et al. 2021).

Urinary creatinine was measured by clinical analyzer and the LOD remained constant at 1 mg/mL (NHANES website). For urinary creatinine below the LOD, we applied 1 mg/mL to the maximal LOD divided by the square root of two. In all analyses, we used urinary creatinine to adjust for urinary dilution (Li et al. 2021). The distributions of the urinary concentrations of OH-PAHs were skewed; and thus, each PAH used in our analysis was transformed by urinary creatinine and log conversion.

### 2.3. Lifestyle index

As done in previous studies (Jin et al. 2021; Li et al. 2018b; Li et al. 2020), a lifestyle index was calculated by combining five components: drinking, smoking, BMI, physical activity, and diet. Each component was assigned a point of 0 or 1; and thus, the lifestyle index ranged from 0 to 5, with higher points indicating a healthier lifestyle. Lifestyle was classified as three categories according to the index: unhealthy (lifestyle index = 0 or 1), middle (lifestyle index = 2 or 3) and healthy (lifestyle index = 4 or 5) in analysis.

The five components were collected by structured questionnaires or physical examination. For drinking, frequency and volume of current alcohol consumption were self-reported. According to the dietary guidelines in the US, we assigned a point of 1 to males who consumed alcohol two drinks or fewer every day and females who consumed alcohol one drink or fewer every day (Zhang et al. 2021); otherwise, we assigned a point of 0. For smoking, we assigned a point of 1 to participants who reported that they smoked less than 100 cigarettes in a lifetime (non-smoking); otherwise, we assigned a point of 0. BMI was calculated as weight in kilograms divided by height in meters squared. We assigned point of 1 to participants whose BMI was higher than 18.5 kg/m^2^ and lower than 25.0 kg/m^2^. For physical activity, we assigned a point of 1 to participants whose metabolic equivalent (MET) score was in the top tercile (Zhang et al. 2021). For diet, an in-person 24-hour dietary recall at the MEC and a follow-up 24-hour dietary recall by phone 3 to 10 days later were conducted by the NHANES interviewer. Dietary quality was assessed using the Healthy Eating Index-2010 (HEI-2010), a validated measurement for how well diets abide by the 2010 Dietary Guidelines for Americans (Bowman SA ; Kirkpatrick et al. 2018). HEI-2010 ranges from 0 to 100, with higher values indicating healthier diets that more closely align with the 2010 Dietary Guidelines for Americans (Kinderknecht et al. 2020). Participants whose HEI-2010 falling into the top two fifths of distribution were assigned a point of 1; otherwise, they were assigned a point of 0 (Li et al. 2018b; Li et al. 2020).

### 2.4. Phenotypic Age Acceleration (PhenoAge.Accel)

We developed PhenoAge based on data from NHANES III (Levine et al. 2018; Liu et al. 2018). PhenoAge was calculated from chronological age (CA) and nine biomarkers (i.e., creatinine, albumin, [log] C-reactive protein, glucose, alkaline phosphatase, lymphocyte percent, red cell distribution width, mean cell volume, and white blood cell count) which had been selected through a Cox Proportional Hazards Elastic Net model for mortality based on ten-fold cross-validation. PhenoAge (unit in years) represents person’s expected age within the population, which is consistent with a person’s estimated hazard of mortality as a function of his/her chemistry biomarkers profile (Levine et al. 2018; Liu et al. 2018). The equation to calculate PhenoAge is shown below:

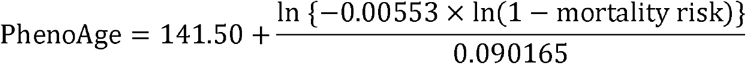

where

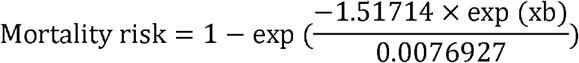

and

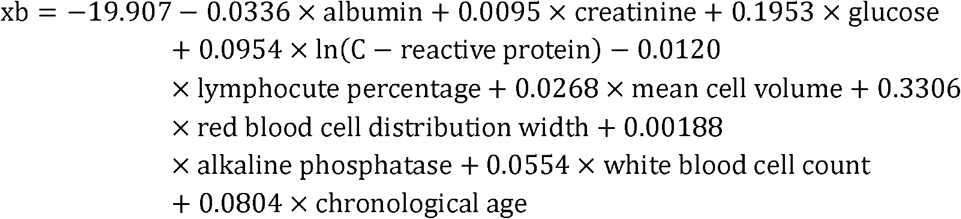

As mentioned above, we calculated a novel aging measure, PhenoAge Acceleration (PhenoAge.Accel), defined as residual from a linear model when regressing PhenoAge on CA. PhenoAge.Accel represents phenotypic aging after accounting for the effect of CA, i.e., a person is phenotypically older (younger) if his/her PhenoAge.Accel>0 (<0) than expected based on his/her CA (Levine et al. 2018; Liu et al. 2018).

### 2.5. Covariates

Obtained through structured questionnaires, our covariates included CA, sex (male and female), race/ethnicity (non-Hispanic White, non-Hispanic Black, Mexican American, and others), education level (less than high school (HS), HS/general educational development (GED), above HS Grad), PIR (< 1, 1-3, ≥ 3) (You et al. 2021).

### 2.6. Statistical analyses

We presented basic characteristics of the participants in total and by sex, using mean ± SD for continuous variables and numbers (percentage) for categorical variables. The correlation between each PAH was analyzed using scatter plot and Pearson’s coefficient matrix.

We used general linear regression models to examine the associations of OH-PAHs and lifestyle with PhenoAge.Accel, respectively. To account for the complex sampling procedure, we considered PAH weight (provided by NHANES) in analyses. We considered two models. In Model 1, we adjusted for CA and sex. In Model 2, we additionally adjusted for race/ethnicity, education level, and PIR. To examine the interaction effect of OH-PAHs (two categories: high and low using median as the cutoff) and lifestyle category (unhealthy, middle, and healthy) on phenotypic aging, we added cross-product terms and tested the statistical significance in Model 1 and 2. To better explicitly illustrate the possible interaction effects, we first examined the associations between OH-PAHs and PhenoAge.Accel, stratified by lifestyle category, and this could suggest the potential modifying effect of lifestyle. Second, we constructed six subgroups by OH-PAHs (two categories) and lifestyle category, and re-examined the associations between OH-PAHs and PhenoAge.Accel, using the group with “unhealthy lifestyle” and “high” OH-PAHs as the reference. We documented the coefficient and corresponding 95% confidence interval (CI).

Given the established sex differences in health effect of ambient air pollution (Shin et al. 2022), we repeated the analyses above stratified by sex. Moreover, to test the robustness of our findings, we performed a sensitivity analysis by replacing the PAH weight with the blood weight (provided by NHANES) and repeated analyses.

All analyses were performed using SAS version 9.4 (SAS Institute, Cary, NC) and R version 4.1.1. Statistically significance was defined as a two-tailed P value < 0.05.

## 3. Results

### 3.1. Basic characteristics of study participants

Table 1 shows the basic characteristics of the participants. The mean CA (± SD) of study participants was 48.2 ± 17.6 years. A total of 1292 participants were female. In all participants, the mean values of ∑NAP, ∑FLU, ∑PHE, ∑PYR, and ∑OH-PAH were 6.5 ± 2.0, 0.9 ± 2.0, -0.6 ± 1.9, -0.3 ± 0.9, and 6.5 ± 5.8 (ng/g Cr), respectively. As shown in Figure S2, a strong correlation between each two of PAHs was revealed. The mean value of PhenoAge (± SD) was 42.5 ± 19.6 years; and PhenoAge.Accel roughly exhibits a normal distribution (Fig. 1).

**Table 1.**
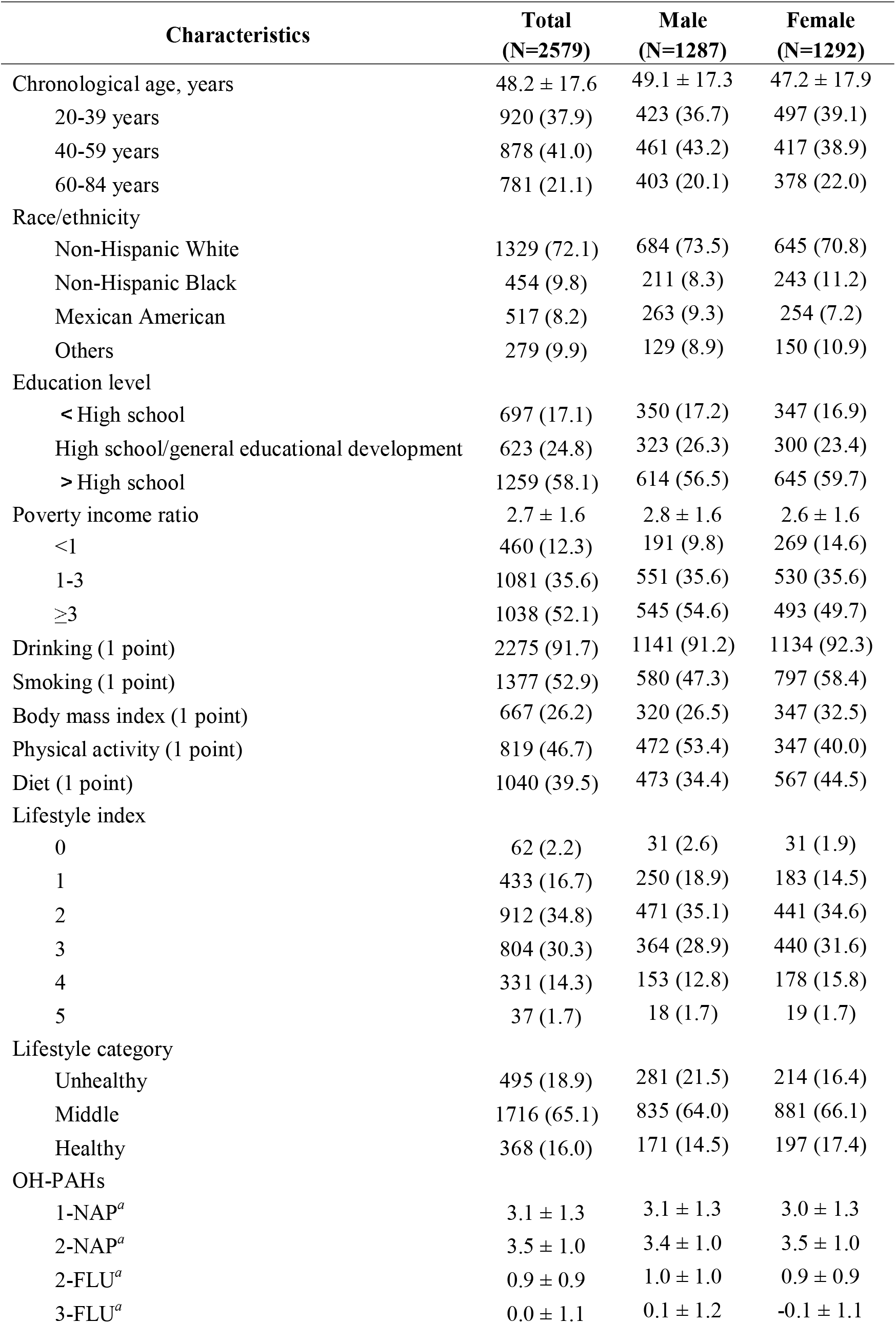
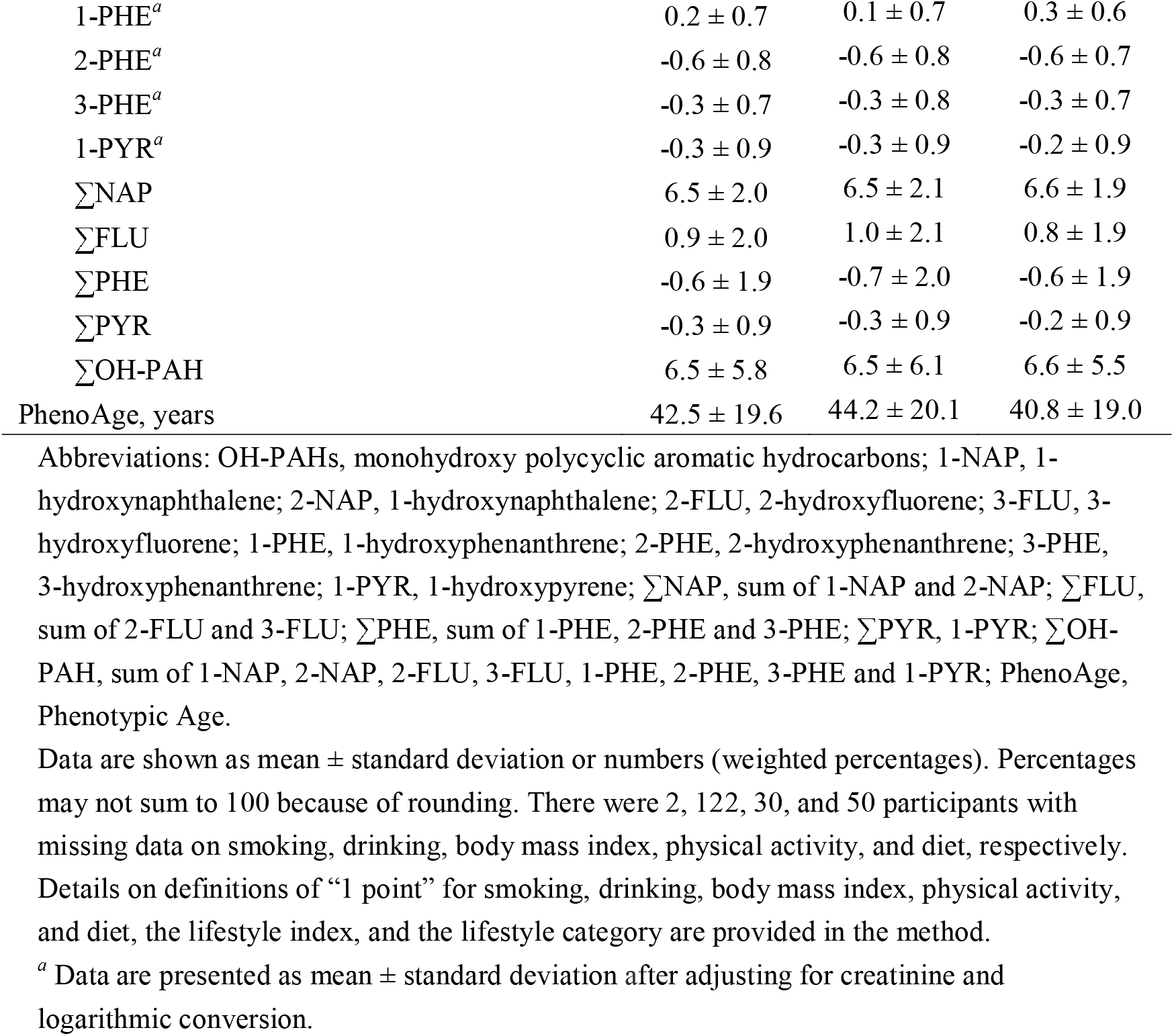
Basic characteristics of study participants in total and by sex.

| Characteristics | Total<br>(N=2579) | Male<br>(N=1287) | Female<br>(N=1292) |
| --- | --- | --- | --- |
| Chronological age, years | 48.2 ± 17.6 | 49.1 ± 17.3 | 47.2 ± 17.9 |
| 20-39 years | 920 (37.9) | 423 (36.7) | 497 (39.1) |
| 40-59 years | 878 (41.0) | 461 (43.2) | 417 (38.9) |
| 60-84 years | 781 (21.1) | 403 (20.1) | 378 (22.0) |
| Race/ethnicity |  |  |  |
| Non-Hispanic White | 1329 (72.1) | 684 (73.5) | 645 (70.8) |
| Non-Hispanic Black | 454 (9.8) | 211 (8.3) | 243 (11.2) |
| Mexican American | 517 (8.2) | 263 (9.3) | 254 (7.2) |
| Others | 279 (9.9) | 129 (8.9) | 150 (10.9) |
| Education level |  |  |  |
| < High school | 697 (17.1) | 350 (17.2) | 347 (16.9) |
| High school/general educational development | 623 (24.8) | 323 (26.3) | 300 (23.4) |
| > High school | 1259 (58.1) | 614 (56.5) | 645 (59.7) |
| Poverty income ratio | 2.7 ± 1.6 | 2.8 ± 1.6 | 2.6 ± 1.6 |
| <1 | 460 (12.3) | 191 (9.8) | 269 (14.6) |
| 1-3 | 1081 (35.6) | 551 (35.6) | 530 (35.6) |
| ≥3 | 1038 (52.1) | 545 (54.6) | 493 (49.7) |
| Drinking (1 point) | 2275 (91.7) | 1141 (91.2) | 1134 (92.3) |
| Smoking (1 point) | 1377 (52.9) | 580 (47.3) | 797 (58.4) |
| Body mass index (1 point) | 667 (26.2) | 320 (26.5) | 347 (32.5) |
| Physical activity (1 point) | 819 (46.7) | 472 (53.4) | 347 (40.0) |
| Diet (1 point) | 1040 (39.5) | 473 (34.4) | 567 (44.5) |
| Lifestyle index |  |  |  |
| 0 | 62 (2.2) | 31 (2.6) | 31 (1.9) |
| 1 | 433 (16.7) | 250 (18.9) | 183 (14.5) |
| 2 | 912 (34.8) | 471 (35.1) | 441 (34.6) |
| 3 | 804 (30.3) | 364 (28.9) | 440 (31.6) |
| 4 | 331 (14.3) | 153 (12.8) | 178 (15.8) |
| 5 | 37 (1.7) | 18 (1.7) | 19 (1.7) |
| Lifestyle category |  |  |  |
| Unhealthy | 495 (18.9) | 281 (21.5) | 214 (16.4) |
| Middle | 1716 (65.1) | 835 (64.0) | 881 (66.1) |
| Healthy | 368 (16.0) | 171 (14.5) | 197 (17.4) |
| OH-PAHs |  |  |  |
| 1-NAP<sup>a</sup> | 3.1 ± 1.3 | 3.1 ± 1.3 | 3.0 ± 1.3 |
| 2-NAP<sup>a</sup> | 3.5 ± 1.0 | 3.4 ± 1.0 | 3.5 ± 1.0 |
| 2-FLU<sup>a</sup> | 0.9 ± 0.9 | 1.0 ± 1.0 | 0.9 ± 0.9 |
| 3-FLU<sup>a</sup> | 0.0 ± 1.1 | 0.1 ± 1.2 | -0.1 ± 1.1 |
| 1-PHE<sup>a</sup> | 0.2 ± 0.7 | 0.1 ± 0.7 | 0.3 ± 0.6 |
| 2-PHE<sup>a</sup> | -0.6 ± 0.8 | -0.6 ± 0.8 | -0.6 ± 0.7 |
| 3-PHE<sup>a</sup> | -0.3 ± 0.7 | -0.3 ± 0.8 | -0.3 ± 0.7 |
| 1-PYR<sup>a</sup> | -0.3 ± 0.9 | -0.3 ± 0.9 | -0.2 ± 0.9 |
| ΣNAP | 6.5 ± 2.0 | 6.5 ± 2.1 | 6.6 ± 1.9 |
| ΣFLU | 0.9 ± 2.0 | 1.0 ± 2.1 | 0.8 ± 1.9 |
| ΣPHE | -0.6 ± 1.9 | -0.7 ± 2.0 | -0.6 ± 1.9 |
| ΣPYR | -0.3 ± 0.9 | -0.3 ± 0.9 | -0.2 ± 0.9 |
| ΣOH-PAH | 6.5 ± 5.8 | 6.5 ± 6.1 | 6.6 ± 5.5 |
| PhenoAge, years | 42.5 ± 19.6 | 44.2 ± 20.1 | 40.8 ± 19.0 |

Abbreviations: OH-PAHs, monohydroxy polycyclic aromatic hydrocarbons; 1-NAP, 1-hydroxynaphthalene; 2-NAP, 1-hydroxynaphthalene; 2-FLU, 2-hydroxyfluorene; 3-FLU, 3-hydroxyfluorene; 1-PHE, 1-hydroxyphenanthrene; 2-PHE, 2-hydroxyphenanthrene; 3-PHE, 3-hydroxyphenanthrene; 1-PYR, 1-hydroxypyrene; ΣNAP, sum of 1-NAP and 2-NAP; ΣFLU, sum of 2-FLU and 3-FLU; ΣPHE, sum of 1-PHE, 2-PHE and 3-PHE; ΣPYR, 1-PYR; ΣOH-PAH, sum of 1-NAP, 2-NAP, 2-FLU, 3-FLU, 1-PHE, 2-PHE, 3-PHE and 1-PYR; PhenoAge, Phenotypic Age.
Data are shown as mean ± standard deviation or numbers (weighted percentages). Percentages may not sum to 100 because of rounding. There were 2, 122, 30, and 50 participants with missing data on smoking, drinking, body mass index, physical activity, and diet, respectively.
Details on definitions of “1 point” for smoking, drinking, body mass index, physical activity, and diet, the lifestyle index, and the lifestyle category are provided in the method.
<sup>a</sup> Data are presented as mean ± standard deviation after adjusting for creatinine and logarithmic conversion.

**Fig. 1.**
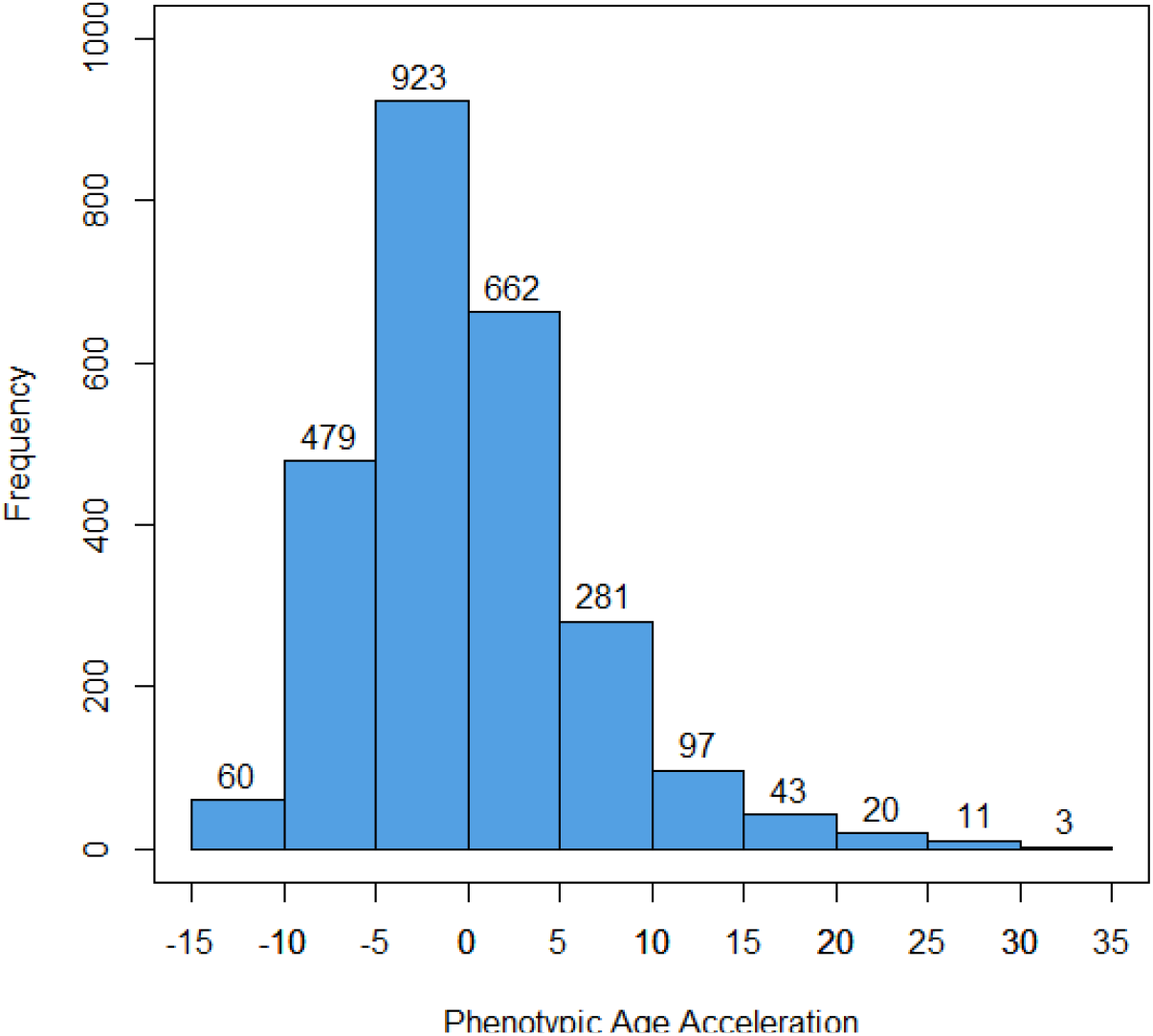
Distribution of participants’ Phenotypic Age Acceleration in this study.

**Fig. 1.**
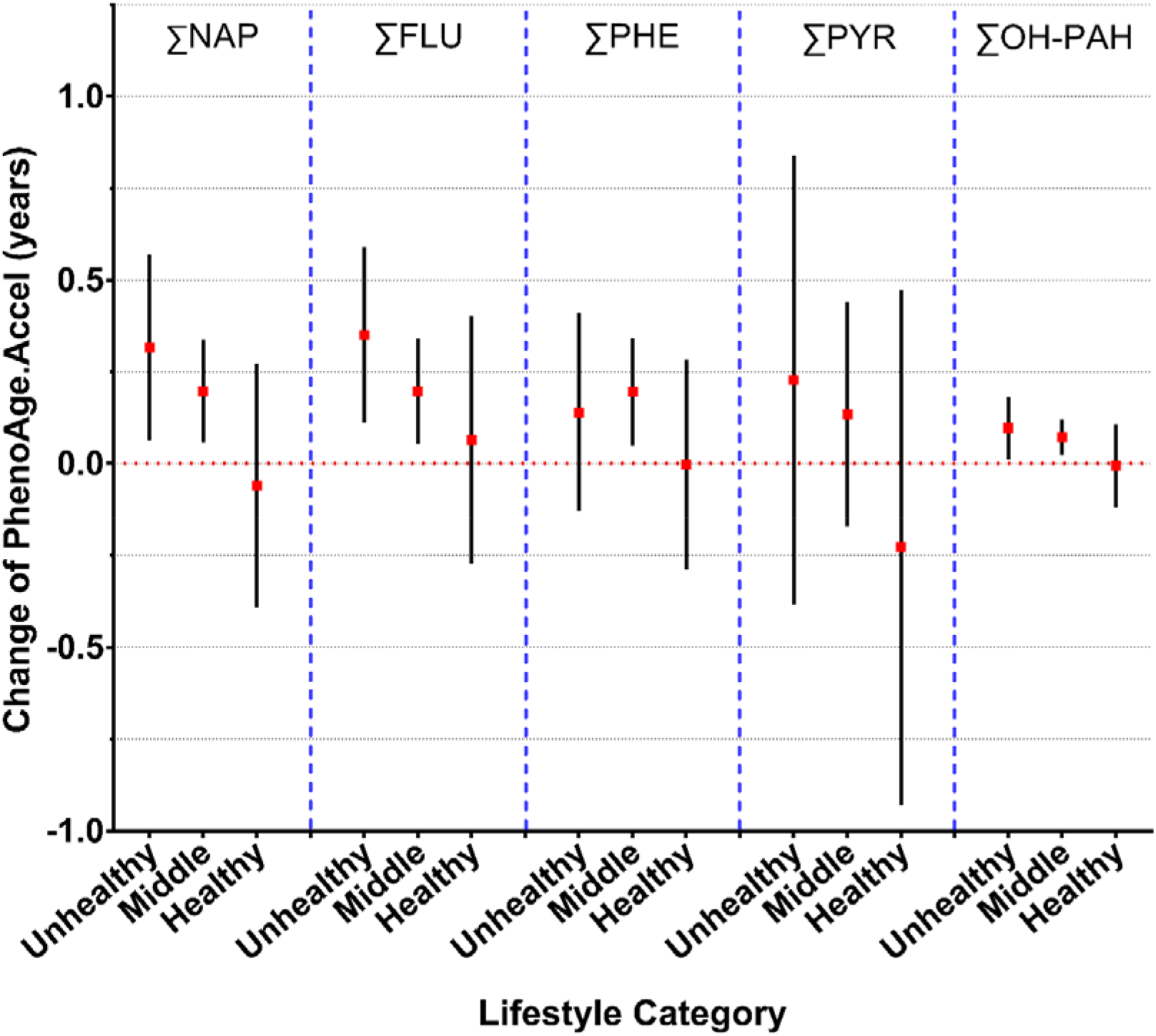
Associations of urinary OH-PAHs with PhenoAge.Accel by lifestyle category. Abbreviations: OH-PAHs, monohydroxy polycyclic aromatic hydrocarbons; ∑NAP, sum of 1-NAP and 2-NAP; ∑FLU, sum of 2-FLU and 3-FLU; ∑PHE, sum of 1-PHE, 2-PHE, and 3-PHE; ∑PYR, 1-PYR; ∑OH-PAH, sum of 1-NAP, 2-NAP, 2-FLU, 3-FLU, 1-PHE, 2-PHE, 3-PHE and 1-PYR; PhenoAge.Accel, Phenotypic Age acceleration.

### 3.2. Associations of OH-PAHs and lifestyle with PhenoAge.Accel

In Table 2, after adjustment for CA and sex (Model 1), we observed that all OH-PAHs were significantly associated with PhenoAge.Accel (all P < 0.05). For instance, one unit increase in ∑NAP was associated with 0.37 year (95% CI: 0.26, 0.48) increase in PhenoAge.Accel and one unit increase in ∑OH-PAH was associated with 0.13 year (95% CI: 0.10, 0.17) increase in PhenoAge.Accel. After further adjusting for race/ethnicity, education level, and PIR in Model 2, the statistically significant associations of OH-PAHs with PhenoAge.Accel were maintained.

**Table 2.** Associations of urinary OH-PAHs and lifestyle with PhenoAge.Accel, respectively.

|  | **Model 1** |  | **Model 2** |  |
| --- | --- | --- | --- | --- |
|  | **β (95%CI)** | **P** | **β (95%CI)** | **P** |
| **OH-PAHs** |  |  |  |  |
| ΣNAP | 0.37 (0.26, 0.48) | <0.001 | 0.30 (0.19, 0.42) | <0.001 |
| ΣFLU | 0.43 (0.32, 0.54) | <0.001 | 0.35 (0.24, 0.46) | <0.001 |
| ΣPHE | 0.26 (0.14, 0.38) | <0.001 | 0.22 (0.11, 0.34) | <0.001 |
| ΣPYR | 0.40 (0.14, 0.66) | 0.002 | 0.26 (0.01, 0.52) | 0.046 |
| ΣOH-PAH | 0.13 (0.09, 0.17) | <0.001 | 0.11 (0.07, 0.15) | <0.001 |
| **Lifestyle category** |  |  |  |  |
| Unhealthy | Reference | — | Reference | — |
| Middle | -1.89 (-2.48, -1.31) | <0.001 | -1.72 (-2.30, -1.14) | <0.001 |
| Healthy | -4.24 (-5.01, -3.48) | <0.001 | -3.80 (-4.56, -3.04) | <0.001 |

Abbreviations: OH-PAHs, monohydroxy polycyclic aromatic hydrocarbons; CI, confidence interval; ΣNAP, sum of 1-NAP and 2-NAP; ΣFLU, sum of 2-FLU and 3-FLU; ΣPHE, sum of 1-PHE, 2-PHE, and 3-PHE; ΣPYR, 1-PYR; ΣOH-PAH, sum of 1-NAP, 2-NAP, 2-FLU, 3-FLU, 1-PHE, 2-PHE, 3-PHE, and 1-PYR; PhenoAge.Accel, Phenotypic Age Acceleration. Each OH-PAH was included into the models separately. Model 1 adjusted for chronological age and sex. Model 2 additionally adjusted for race/ethnicity, education level, and poverty income ratio based on Model 1.

As shown in Table 2, compared to participants with an unhealthy lifestyle, those who with a middle lifestyle (β = -1.89, 95% CI: -2.48, -1.30) and a healthy lifestyle (β = -4.24, 95% CI: -5.00, -3.48) had lower level of PhenoAge.Accel. Similar results were observed in Model 2.

### 3.3. The interaction effects of OH-PAHs and lifestyle on PhenoAge.Accel

We did not observe statistically significant interactions of OH-PAHs and lifestyle on PhenoAge.Accel (data not shown). However, our results suggested that lifestyle might attenuate the effects of OH-PAHs on PhenoAge.Accel. As shown in Figure 2, the association between ∑OH-PAH and PhenoAge.Accel was statistically significant (β = 0.10, P = 0.026) among participants with an unhealthy lifestyle, but the association was non-significant (β = -0.01, P = 0.917) for participants with a healthy lifestyle.

**Fig. 2.**
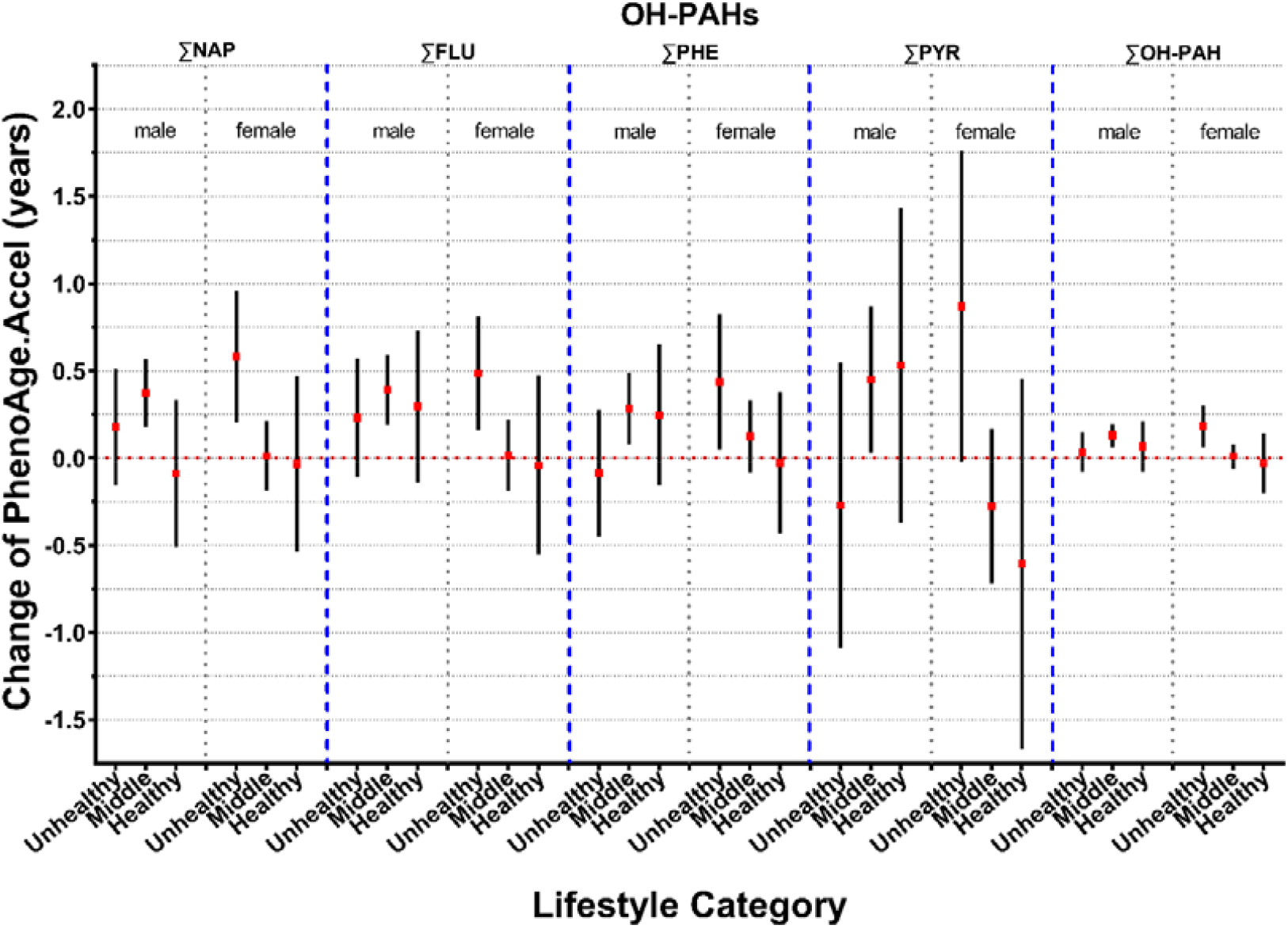
Associations of OH-PAHs with PhenoAge.Accel by sex and lifestyle category. Abbreviations: OH-PAHs, monohydroxy polycyclic aromatic hydrocarbons; ∑NAP, sum of 1-NAP and 2-NAP; ∑FLU, sum of 2-FLU and 3-FLU; ∑PHE, sum of 1-PHE, 2-PHE, and 3-PHE; ∑PYR, 1-PYR; ∑OH-PAH, sum of 1-NAP, 2-NAP, 2-FLU, 3-FLU, 1-PHE, 2-PHE, 3-PHE and 1-PYR; PhenoAge.Accel, Phenotypic Age Acceleration.

Among six subgroups constructed by OH-PAHs (by median, two categories) and lifestyle (three categories), we observed that subgroups with a healthy lifestyle had the lowest level of PhenoAge.Accel regardless of a high or low PAH (Figure S3).

Results were from a model adjusted for chronological age, sex, race/ethnicity, education level, and poverty income ratio.

### 3.4. Additional analysis

Table S1 shows the significant associations of OH-PAHs and lifestyle with PhenoAge.Accel in males and females. For instance, one unit increase in ∑NAP was associated with 0.44 year (95% CI: 0.28, 0.60) increase in PhenoAge.Accel among males and 0.32 year (95% CI: 0.16, 0.31) increase in PhenoAge.Accel among females. Interestingly, we observed statistically significant interactions between several OH-PAHs and lifestyle (e.g., P for interaction = 0.029 for ∑NAP * lifestyle category) on PhenoAge.Accel among females (Table S2). As shown in Figure 3, stratified by lifestyle, for females with an unhealthy lifestyle, we observed a significant association between OH-PAHs and PhenoAge while we did not observe similar associations for females with a middle or healthy lifestyle. In sensitivity analyses, we replaced the PAH weight with the blood weight and found that the results were maintained when: (1) re-examining the associations of urinary OH-PAHs and lifestyle with PhenoAge.Accel (Table S3); (2) re-examining the associations of urinary OH-PAHs and lifestyle with PhenoAge.Accel stratified by sex (Table S4); (3) re-examining the interaction between OH-PAHs and lifestyle stratified by sex (Table S5); (4) re-examining the associations of urinary OH-PAHs with PhenoAge.Accel by lifestyle category (Figure S4); (5) re-examining the associations of urinary OH-PAHs with PhenoAge.Accel by sex and lifestyle category (Figure S5); and (6) re-examining the joint associations of OH-PAHs and lifestyle with PhenoAge.Accel (Figure S6).

Results were from a model adjusted for chronological age, sex, race/ethnicity, education level, and poverty income ratio.

## 4. Discussion

Based on the data from NHANES 2001-2010, we found that OH-PAHs and lifestyle were independently associated with PhenoAge.Accel. We also found interaction effects of several OH-PAHs and lifestyle on PhenoAge.Accel among females. Since accelerated aging indicates adverse health issues in later life, our findings highlight the importance of adherence to a healthy lifestyle, particularly for females who are exposed to high PAHs.

For the first time, we demonstrated that high PAHs were associated with accelerated phenotypic aging. Qiao et al (Qiao et al. 2017) revealed the adverse effect of PAHs on skin aging. Pavanello et al (Pavanello et al. 2020) suggested that certain preventable everyday life exposures to PAHs reduce leucocyte telomere length and mitochondrial DNA copy number, which were associated with increased risks of mortality and age-related diseases. Studies have reported that exposure to PAHs was associated with accelerations of DNA methylation age (Li et al. 2018a). Though our findings complement these results to some extent, PhenoAge.Accel represents a multisystem aging process, with complex underlying biological mechanisms. Our findings may provide an exciting new avenue for investigating the underlying biological mechanisms of how air pollutions induce aging-related adverse health outcomes (including age-related diseases and mortality) through aging.

The detrimental effects of exposures to PAHs on phenotypic aging were attenuated by a healthy lifestyle in females, which has not been reported previously. Although some results did not reach the statistical significance, the trend was observed when examining the association of PAHs with phenotypic aging stratified by sex (Fig. 2). For example, detrimental effects of PAHs on phenotypic aging in a subgroup with an unhealthy lifestyle were not observed in a subgroup with a healthy lifestyle. The findings have important public implications, suggesting adaption to a healthy lifestyle to attenuate the detrimental effect of exposure to air pollutions (e.g., PAHs) on phenotypic aging among females. However, chance findings cannot be rule out when interpreting the results given the relatively small sample size of the stratified analyses by sex.

One plausible underlying biological mechanism of the interesting interaction effect of PAHs and lifestyle on phenotypic aging is related to oxidative stress and inflammation. Higher exposure to PAHs could result in increased oxidative stress (Zhu et al. 2021) and oxidative DNA damage (Kuang et al. 2013), two key determinants of aging process. PAHs can induce inflammatory response in the body including elevating high C-reactive protein levels (a marker reflecting the systemic inflammation and was included in calculation of PhenoAge) (Farzan et al. 2016). Interestingly, many components included in our lifestyle index have been demonstrated to be associated with inflammation or oxidative stress as well, such as diet quality (de Castro Mendes et al. 2020), physical activity (Tu et al. 2020) and BMI. For instance, adipose tissue can produce proinflammatory cytokines (TNF-α, IL-1β and IL-6) related to persistent inflammation and oxidative stress, and adipokines (leptin, adiponectin, plasminogen activator inhibitor, and macrophage migration inhibitory factor) which mediate the inflammatory response (Fain 2006). Building on the above evidence, we speculate that high PAHs were associated with accelerated phenotypic aging, partially through increased oxidative stress and inflammation.

There are several strengths in this study. First, NHANES employs thorough quality control and quality assurance procedures to ensure the validity of the data. Second, we concentrated on a lifestyle index rather than a single component, to comprehensively capture the complex relations of many lifestyle factors. Third, the study outcome, PhenoAge.Accel, is not simply a measure of mortality; instead, it may be a marker that could track the aging process before diseases become clinically evident. Thus, the findings have more public health implications in terms of disease preventions in community settings.

However, this study has some limitations. First, information on several lifestyle components was self-reported; and thus, recall bias is possible. Second, the level of PAHs in spot urine was performed by a single measurement; and thus, intra-person variability and long-term average exposure may not be reflected. Third, the temporal relationships between PAHs, lifestyle and phenotypic aging cannot be determined given the cross-sectional study design. Fourth, the analytic sample was selected based on multiple criteria such as with information on both PAHs and PhenoAge; and thus, was relatively small (for stratification analysis) and might be biased to some extent. Finally, although we included many covariates, there may still be residual and unmeasured confounders.

## 5. Conclusions

In this sample of an US population, for the first time, we demonstrated that both OH-PAHs and lifestyle were independently associated with phenotypic aging and there were statistically significant interactions between OH-PAHs and lifestyle on phenotypic aging among females. The findings provide important insights into the underlying biological mechanism of how exposure to PAHs and lifestyle accelerate aging. Furthermore, the findings highlight the importance of adherence to a healthy lifestyle to attenuate accelerated phenotypic aging, which has significant public health implications.

## Data Availability

All data produced in the present study are available upon reasonable request to the authors.

## Abbreviations

OH-PAHs: monohydroxy polycyclic aromatic hydrocarbons
PhenoAge.Accel: Phenotypic Age Acceleration
CI: confidence interval
∑NAP: Naphthalene, sum of 1- and 2-hydroxynaphthalene
∑FLU: Fluorene, sum of 2- and 3-hydroxyfluorene
∑PHE: Phenanthrene, sum of 1-, 2-, and 3-hydroxyphenanthrene
∑PYR: Pyrene, 1-hydroxpyrene
∑OH-PAHs: sum of 1-, 2-hydroxynaphthalene, 2-, 3-hydroxyfluorene, 1-, 2-, 3-hydroxyphenanthrene and 1-hydroxpyrene
BMI: body mass index
HNANES: National Health and Nutritional Examination Surveys
MEC: Mobile Exam Center
CDC: Centers for Disease Control and Prevention
PIR: poverty income ratio
Cr: creatinine
SD: standard deviation
LOD: limit of detection
MET: metabolic equivalent
HEI: Healthy Eating Index
CA: chronological age
HS: high school
GED: general educational development

## Acknowledgments

This study was supported by a grant from the National Natural Science Foundation of China (82171584), the 2020 Milstein Medical Asian American Partnership Foundation Irma and Paul Milstein Program for Senior Health project award (ZL), the Fundamental Research Funds for the Central Universities (ZL), a project from the Natural Science Foundation of Zhejiang Province (LQ21H260003), and Zhejiang University Global Partnership Fund (188170-11103). The funders had no role in the study design; data collection, analysis, or interpretation; in the writing of the report; or in the decision to submit the article for publication.

## Reference

SAS code. Division of Cancer Control and Population Science website. https://epi.grants.cancer.gov/hei/sas-code.html (accessed

Abdel-Shafy, H.I.; Mansour, M.S.M. A review on polycyclic aromatic hydrocarbons: Source, environmental impact, effect on human health and remediation. Egyptian Journal of Petroleum 2016;25:107–123. https://doi.org/https://doi.org/10.1016/j.ejpe.2015.03.011

Astuti, Y.; Wardhana, A.; Watkins, J.; Wulaningsih, W. Cigarette smoking and telomere length: A systematic review of 84 studies and meta-analysis. Environ Res 2017;158:480–489. https://doi.org/10.1016/j.envres.2017.06.038

Bowman SA C.J., Friday JE, Thoerig RC, Moshfegh AJ. Food Patterns Equivalents Database 2005-06: Methodology and User Guide 2014. https://www.ars.usda.gov/ARSUserFiles/80400530/pdf/fped/FPED_0506.pdf (accessed

Brockmeyer, T.; Friederich, H.C.; Schmidt, U. Advances in the treatment of anorexia nervosa: a review of established and emerging interventions. Psychol Med 2018;48:1228–1256. https://doi.org/10.1017/S0033291717002604

Buckley, T.J.; Lioy, P.J. An examination of the time course from human dietary exposure to polycyclic aromatic hydrocarbons to urinary elimination of 1-hydroxypyrene. British Journal of Industrial Medicine 1992;49:113. https://doi.org/10.1136/oem.49.2.113

Burstyn, I.; Kromhout, H.; Partanen, T.; Svane, O.; Langård, S.; Ahrens, W.; Kauppinen, T.; Stücker, I.; Shaham, J.; Heederik, D.; Ferro, G.; Heikkilä, P.; Hooiveld, M.; Johansen, C.; Randem, B.G.; Boffetta, P. Polycyclic aromatic hydrocarbons and fatal ischemic heart disease. Epidemiology 2005;16:744–750. https://doi.org/10.1097/01.ede.0000181310.65043.2f

de Castro Mendes, F.; Paciência, I.; Cavaleiro Rufo, J.; Silva, D.; Cunha, P.; Farraia, M.; Delgado, L.; Garcia-Larsen, V.; Severo, M.; Moreira, A.; Moreira, P. The inflammatory potential of diet impacts the association between air pollution and childhood asthma. Pediatric Allergy and Immunology 2020;31:290–296. https://doi.org/https://doi.org/10.1111/pai.13185

Fain, J.N. Release of interleukins and other inflammatory cytokines by human adipose tissue is enhanced in obesity and primarily due to the nonfat cells. Vitam Horm 2006;74:443–477. https://doi.org/10.1016/s0083-6729(06)74018-3

Fan, J.; Cai, S.; Mi, S.; Chen, H.; Chen, D.; Fan, C.; Sun, L.; Li, Y. Association of urinary phthalate metabolites with osteoarthritis in American adults: Results from the national health and nutrition examination survey 2003–2014. Chemosphere 2021;268:128807. https://doi.org/https://doi.org/10.1016/j.chemosphere.2020.128807

Farzan, S.F.; Chen, Y.; Trachtman, H.; Trasande, L. Urinary polycyclic aromatic hydrocarbons and measures of oxidative stress, inflammation and renal function in adolescents: NHANES 2003–2008. Environmental Research 2016;144:149–157. https://doi.org/https://doi.org/10.1016/j.envres.2015.11.012

Ferrucci, L.; Levine, M.E.; Kuo, P.L.; Simonsick, E.M. Time and the Metrics of Aging. Circ Res 2018;123:740–744. https://doi.org/10.1161/circresaha.118.312816

Grainger, J.; Huang, W.; Patterson, D.G.; Turner, W.E.; Pirkle, J.; Caudill, S.P.; Wang, R.Y.; Needham, L.L.; Sampson, E.J. Reference range levels of polycyclic aromatic hydrocarbons in the US population by measurement of urinary monohydroxy metabolites. Environmental Research 2006;100:394–423. https://doi.org/https://doi.org/10.1016/j.envres.2005.06.004

Jin, S.; Li, C.; Cao, X.; Chen, C.; Ye, Z.; Liu, Z. Association of lifestyle with mortality and the mediating role of aging among older adults in China. Arch Gerontol Geriatr 2021;98:104559. https://doi.org/10.1016/j.archger.2021.104559

Kennedy, B.K.; Berger, S.L.; Brunet, A.; Campisi, J.; Cuervo, A.M.; Epel, E.S.; Franceschi, C.; Lithgow, G.J.; Morimoto, R.I.; Pessin, J.E.; Rando, T.A.; Richardson, A.; Schadt, E.E.; Wyss-Coray, T.; Sierra, F. Geroscience: Linking Aging to Chronic Disease. Cell 2014;159:709–713. https://doi.org/https://doi.org/10.1016/j.cell.2014.10.039

Kim, K.-H.; Jahan, S.A.; Kabir, E.; Brown, R.J.C. A review of airborne polycyclic aromatic hydrocarbons (PAHs) and their human health effects. Environment International 2013;60:71–80. https://doi.org/https://doi.org/10.1016/j.envint.2013.07.019

Kinderknecht, K.; Harris, C.; Jones-Smith, J. Association of the Healthy, Hunger-Free Kids Act With Dietary Quality Among Children in the US National School Lunch Program. JAMA 2020;324:359–368. https://doi.org/10.1001/jama.2020.9517

Kirkpatrick, S.I.; Reedy, J.; Krebs-Smith, S.M.; Pannucci, T.E.; Subar, A.F.; Wilson, M.M.; Lerman, J.L.; Tooze, J.A. Applications of the Healthy Eating Index for Surveillance, Epidemiology, and Intervention Research: Considerations and Caveats. Journal of the Academy of Nutrition and Dietetics 2018;118:1603–1621. https://doi.org/https://doi.org/10.1016/j.jand.2018.05.020

Kuang, D.; Zhang, W.; Deng, Q.; Zhang, X.; Huang, K.; Guan, L.; Hu, D.; Wu, T.; Guo, H. Dose-Response Relationships of Polycyclic Aromatic Hydrocarbons Exposure and Oxidative Damage to DNA and Lipid in Coke Oven Workers. Environmental Science & Technology 2013;47:7446–7456. https://doi.org/10.1021/es401639x

Levine, M.E.; Lu, A.T.; Quach, A.; Chen, B.H.; Assimes, T.L.; Bandinelli, S.; Hou, L.; Baccarelli, A.A.; Stewart, J.D.; Li, Y.; Whitsel, E.A.; Wilson, J.G.; Reiner, A.P.; Aviv, A.; Lohman, K.; Liu, Y.; Ferrucci, L.; Horvath, S. An epigenetic biomarker of aging for lifespan and healthspan. Aging (Albany NY) 2018;10:573–591. https://doi.org/10.18632/aging.101414

Li, J.; Li, X.; Xia, Y.; Fan, H.; Fan, D.; Xi, X.; Ye, Q.; Zhu, Y.; Xiao, C. Subgroup analysis of the relationship between polycyclic aromatic hydrocarbons and rheumatoid arthritis: Data from the National Health and Nutrition Examination Survey, 2003-2014. Sci Total Environ 2021;775:145841. https://doi.org/10.1016/j.scitotenv.2021.145841

Li, J.; Zhu, X.; Yu, K.; Jiang, H.; Zhang, Y.; Wang, B.; Liu, X.; Deng, S.; Hu, J.; Deng, Q.; Sun, H.; Guo, H.; Zhang, X.; Chen, W.; Yuan, J.; He, M.; Bai, Y.; Han, X.; Liu, B.; Liu, C.; Guo, Y.; Zhang, B.; Zhang, Z.; Hu, F.B.; Gao, W.; Li, L.; Lathrop, M.; Laprise, C.; Liang, L.; Wu, T. Exposure to Polycyclic Aromatic Hydrocarbons and Accelerated DNA Methylation Aging. Environmental Health Perspectives 2018a;126:067005. https://doi.org/doi:10.1289/EHP2773

Li, Y.; Pan, A.; Wang, D.D.; Liu, X.; Dhana, K.; Franco, O.H.; Kaptoge, S.; Di Angelantonio, E.; Stampfer, M.; Willett, W.C.; Hu, F.B. Impact of Healthy Lifestyle Factors on Life Expectancies in the US Population. Circulation 2018b;138:345–355. https://doi.org/10.1161/circulationaha.117.032047

Li, Y.; Schoufour, J.; Wang, D.D.; Dhana, K.; Pan, A.; Liu, X.; Song, M.; Liu, G.; Shin, H.J.; Sun, Q.; Al-Shaar, L.; Wang, M.; Rimm, E.B.; Hertzmark, E.; Stampfer, M.J.; Willett, W.C.; Franco, O.H.; Hu, F.B. Healthy lifestyle and life expectancy free of cancer, cardiovascular disease, and type 2 diabetes: prospective cohort study. BMJ 2020;368:l6669. https://doi.org/10.1136/bmj.l6669

Liu, Z.; Kuo, P.L.; Horvath, S.; Crimmins, E.; Ferrucci, L.; Levine, M. A new aging measure captures morbidity and mortality risk across diverse subpopulations from NHANES IV: A cohort study. PLoS Med 2018;15:e1002718. https://doi.org/10.1371/journal.pmed.1002718

McClean, M.D.; Rinehart, R.D.; Ngo, L.; Eisen, E.A.; Kelsey, K.T.; Herrick, R.F. Inhalation and dermal exposure among asphalt paving workers. Ann Occup Hyg 2004;48:663–671. https://doi.org/10.1093/annhyg/meh062

Motorykin, O.; Santiago-Delgado, L.; Rohlman, D.; Schrlau, J.E.; Harper, B.; Harris, S.; Harding, A.; Kile, M.L.; Massey Simonich, S.L. Metabolism and excretion rates of parent and hydroxy-PAHs in urine collected after consumption of traditionally smoked salmon for Native American volunteers. Science of The Total Environment 2015;514:170–177. https://doi.org/https://doi.org/10.1016/j.scitotenv.2015.01.083

Pavanello, S.; Campisi, M.; Mastrangelo, G.; Hoxha, M.; Bollati, V. The effects of everyday-life exposure to polycyclic aromatic hydrocarbons on biological age indicators. Environ Health 2020;19:128. https://doi.org/10.1186/s12940-020-00669-9

Qiao, Y.; Li, Q.; Du, H.-Y.; Wang, Q.-W.; Huang, Y.; Liu, W. Airborne polycyclic aromatic hydrocarbons trigger human skin cells aging through aryl hydrocarbon receptor. Biochemical and Biophysical Research Communications 2017;488:445–452. https://doi.org/https://doi.org/10.1016/j.bbrc.2017.04.160

Robinson, O.; Chadeau Hyam, M.; Karaman, I.; Climaco Pinto, R.; Ala-Korpela, M.; Handakas, E.; Fiorito, G.; Gao, H.; Heard, A.; Jarvelin, M.R.; Lewis, M.; Pazoki, R.; Polidoro, S.; Tzoulaki, I.; Wielscher, M.; Elliott, P.; Vineis, P. Determinants of accelerated metabolomic and epigenetic aging in a UK cohort. Aging Cell 2020;19:e13149. https://doi.org/10.1111/acel.13149

Shin, H.H.; Maquiling, A.; Thomson, E.M.; Park, I.W.; Stieb, D.M.; Dehghani, P. Sex-difference in air pollution-related acute circulatory and respiratory mortality and hospitalization. Sci Total Environ 2022;806:150515. https://doi.org/10.1016/j.scitotenv.2021.150515

Stallings-Smith, S.; Mease, A.; Johnson, T.M.; Arikawa, A.Y. Exploring the association between polycyclic aromatic hydrocarbons and diabetes among adults in the United States. Environmental Research 2018;166:588–594. https://doi.org/https://doi.org/10.1016/j.envres.2018.06.041

Tu, R.; Hou, J.; Liu, X.; Li, R.; Dong, X.; Pan, M.; Mao, Z.; Huo, W.; Chen, G.; Guo, Y.; Li, S.; Wang, C. Physical activity attenuated association of air pollution with estimated 10-year atherosclerotic cardiovascular disease risk in a large rural Chinese adult population: A cross-sectional study. Environ Int 2020;140:105819. https://doi.org/10.1016/j.envint.2020.105819

Veyrand, B.; Sirot, V.; Durand, S.; Pollono, C.; Marchand, P.; Dervilly-Pinel, G.; Tard, A.; Leblanc, J.C.; Le Bizec, B. Human dietary exposure to polycyclic aromatic hydrocarbons: results of the second French Total Diet Study. Environ Int 2013;54:11–17. https://doi.org/10.1016/j.envint.2012.12.011

Wang, L.; Hou, J.; Hu, C.; Zhou, Y.; Sun, H.; Zhang, J.; Li, T.; Gao, E.; Wang, G.; Chen, W.; Yuan, J. Mediating factors explaining the associations between polycyclic aromatic hydrocarbons exposure, low socioeconomic status and diabetes: A structural equation modeling approach. Science of The Total Environment 2019;648:1476–1483. https://doi.org/https://doi.org/10.1016/j.scitotenv.2018.08.255

You, Y.; Luo, J.; Elstgeest, L.E.M.; Zhang, Y.; Tan, S.S.; Raat, H. Associations between health indicators and sleep duration of American adults: NHANES 2011-16. Eur J Public Health 2021;31:1204–1210. https://doi.org/10.1093/eurpub/ckab172

Zhang, Y.; Ding, J.; Shen, G.; Zhong, J.; Wang, C.; Wei, S.; Chen, C.; Chen, Y.; Lu, Y.; Shen, H.; Li, W.; Huang, Y.; Chen, H.; Su, S.; Lin, N.; Wang, X.; Liu, W.; Tao, S. Dietary and inhalation exposure to polycyclic aromatic hydrocarbons and urinary excretion of monohydroxy metabolites--a controlled case study in Beijing, China. Environ Pollut 2014;184:515–522. https://doi.org/10.1016/j.envpol.2013.10.005

Zhang, Y.B.; Chen, C.; Pan, X.F.; Guo, J.; Li, Y.; Franco, O.H.; Liu, G.; Pan, A. Associations of healthy lifestyle and socioeconomic status with mortality and incident cardiovascular disease: two prospective cohort studies. BMJ 2021;373:n604. https://doi.org/10.1136/bmj.n604

Zhu, H.; Martinez-Moral, M.P.; Kannan, K. Variability in urinary biomarkers of human exposure to polycyclic aromatic hydrocarbons and its association with oxidative stress. Environ Int 2021;156:106720. https://doi.org/10.1016/j.envint.2021.106720

